# General and Anxiety-Linked Influence of Acute Serotonin Reuptake Inhibition on Neural Responses Associated with Attended Visceral Sensation

**DOI:** 10.1101/2024.01.23.24301647

**Authors:** James J A Livermore, Lina I Skora, Kristian Adamatzky, Sarah N Garfinkel, Hugo D Critchley, Daniel Campbell-Meiklejohn

**Affiliations:** School of Psychology, University of Sussex, Brighton, UK; Donders Institute for Brain, Cognition and Behaviour, Radboud University, Nijmegen, Netherlands; Heinrich Heine Universität, Düsseldorf, Germany; Sussex Centre for Consciousness Science, University of Sussex, Brighton, UK; Brighton and Sussex Medical School, Brighton, UK; Institute of Cognitive Neuroscience, University College London; Sussex Partnership NHS Foundation Trust, UK

**Keywords:** Interoception, Serotonin, Anxiety

## Abstract

Serotonin is known to have state-dependent modulatory influences on exteroceptive sensory processes and the processing of pain, but much less is known about its role in ordinary interoceptive processes and their relationships to affective states. This experiment compared the impact of a selective serotonin reuptake inhibitor (SSRI) (20mg CITALOPRAM), acutely increasing extracellular serotonin, to that of a PLACEBO on the neural processing of ordinary interoceptive sensations and the relationship of these influences to anxious states. Twenty-one healthy young volunteers completed the visceral interoceptive attention (VIA) task with each treatment, focusing attention on heart, stomach, or visual sensation control while scanned with functional magnetic resonance imaging (fMRI). The relative neural interoceptive response (IR) to heart sensation [heart *minus* visual] and stomach sensation [stomach *minus* visual] were compared between treatment conditions, controlling for general effects on sensory processing. CITALOPRAM reduced interoceptive processing in viscerosensory (bilateral posterior insular cortex, stomach-IR) and integrative/affective components (bilateral amygdala, stomach-IR and heart-IR) of known interoceptive pathways. We then searched for state-dependent modulatory effects of CITALOPRAM that varied with current levels of anxiety. The anterior insular cortex response to heart sensation increased with anxiety, replicating prior findings. This relationship disappeared on CITALOPRAM. Preliminary *post hoc* exploration found that CITALOPRAM’S effects on amygdalae response to stomach sensation predicted acute increases and decreases in anxiety. Overall, this evidence of general and state-dependent serotonergic influence advances our understanding of interoception, its regulation, and its relationship to anxious states.

## Main Text

If asked how one feels, it is natural to turn attention to the body. This involves ordinary interoception – the sensing and processing of internal physiological states ^1–3^. The influence of interoception ranges from regulatory reflexes that maintain homeostasis to conscious experiences such as hunger, arousal, emotion, anxiety, and self ^2, 4–7^. Yet, little is known about the involvement of neurotransmitters in these influences, including those influences involved in mental health disorders and their treatment ^8^. Here, we studied the relationship of interoception to acute changes in serotonin and how this association relates to the experience of anxious states.

Serotonin is known to acutely modulate exteroceptive (e.g. auditory, visual) sensory processing according to animal needs and environment – with a complex but overall effect attenuating the central nervous system’s sensory response (e.g. for reviews ^9, 10^). There is good reason to expect similar serotonergic influences for ordinary interoception. For instance, the impacts of acute serotonergic changes on behavioural and neural responses to aversive, exogenous digestive tract stimulation could potentially extend to the modulation of ordinary interoception. In the limited reports available, the effects of acute serotonin manipulations depended on the location of the stimulation, the degree of pain, and the baseline sensitivity of the participant. But overall, while 5HT_3A_ antagonists and single SSRI doses (which increase extracellular serotonin) tend to reduce affective and neural responses to aversive interoceptive sensation, depleting serotonin’s precursor by acute tryptophan depletion (ATD) tends to increase them ^11–16^. Even less is understood about serotonergic influence on interoception of the heart, which can influence affective cognition, such as the detection and memory of fear ^17, 18^. Patients with major depression, generally associated with blunted neural interoceptive responses ^19^, seem to experience higher subjective intensity of heart (and stomach) sensations if chronically treated with SSRIs, but a significant neural correlate of this effect was not reported ^20^. In another study, we found that a single SSRI dose in healthy volunteers increases metacognitive insight into one’s ability to feel one’s heart ^21^. Thus far, inferences about the relationship of serotonin to ordinary interoceptive processing have been limited by the testing of unusual (painful) visceral sensations, the study of patients with disturbed interoception, absent exteroceptive control conditions, the examination of only a single interoceptive domain and/or a lack of neural insight.

An association between serotonin and interoception can also be hypothesised from their shared relationships to anxiety. The *serotonin*-*anxiety* link is demonstrated by considerable preclinical research (e.g. ^22–24^) and knowledge that SSRIs are effective first-line pharmacological agents for treating panic and generalised anxiety disorder ^25, 26^ but can also unpredictably increase anxiety in the short term ^27, 28^. The *interoception*-*anxiety* link is found in the theoretical understanding of the role of interoception in the pathoaetiology and treatment of somatic anxiety symptoms, backed by empirical observations of interoceptive disturbance in the same anxiety disorders that are treated with SSRIs ^7, 29–31^. Modern conceptualisations of irritable bowel syndrome also acknowledge the neural, genetic, homeostatic and pharmacological overlap between gastrointestinal sensation and anxious states ^32^. In the lab, the anxiety-linked behaviours acutely affected by SSRIs can also vary with interoceptive signalling. These include startle (e.g. ^33–36^), fear (e.g. ^17, 37^), and emotion recognition ^38–41^. The neural substrates of interoception, anxiety, and serotonergic impact on cognition also overlap. At serotonin terminals, this occurs within the insular cortex and amygdala ^3, 23, 32, 41–50^. However, serotonin could also influence interoception at serotonin cell bodies of the raphe nuclei. The raphe nuclei contain cells that modulate appetitive, cardiac, respiratory, sensory, and thermal regulatory processes in response to interoceptive information ^51, 52^ and are proposed to play a key role in regulating anxiety. SSRIs disproportionately increase extracellular serotonin at the raphe nuclei following acute doses ^53^. Besides anxiety, SSRIs also often cause emotional blunting, which could also be attributable to the suppressed intensity of interoception or its reduced association with the experience of arousal ^54–56^. Critically, however, all this evidence for serotonergic modulation of normal interoception is indirect – no direct link has yet been made ^57^.

The effects of serotonin on interoception may be state-dependent. Serotonergic effects on exteroceptive processing are sensitive to states of the environment and individual ^10, 58, 59^, suggesting that serotonin may tune the modulatory impact of these states ^60^. Similarly, the effects of serotonin on gastrointestinal interoception have depended on existing sensitivity and the intensity of aversive stimulation ^11, 12, 15^. Anxiety is a state that is self-evidently linked to interoception. Correspondingly, the anterior insular cortex responds more during focus on cardiac interoception in anxious individuals and anxious individuals can be more susceptible to interoceptive influences on cognition ^18, 43, 44^. We therefore hypothesised that some influences of serotonin on interoception might only be observed during anxious states, potentially modulating anxiety’s relationship to interoception. Selectivity of serotonin effects for anxious states has been previously observed within the amygdala, wherein a single SSRI dose reduced the behavioural and amygdala neural response to observed fearful faces in patients with depression, in whom these responses were initially increased and who were also experiencing increased anxiety, but not in healthy controls ^61, 62^.

Neural responses to normal physiological sensations can be probed by asking individuals to shift and maintain attention to a visceral sensation, which typically enhances the regional neural response underlying the corresponding interoceptive representation ^20, 49, 63–66^. We combined this approach with the established pharmacological technique of using a single SSRI dose in healthy volunteers to test the influence of increased extracellular serotonin ^41, 50^. While long-term SSRI doses are essential for modelling the mechanisms of their delayed therapeutic effects, acute dose studies provide insight into the impacts of immediate serotonin changes on cognition without confounds of receptor desensitisation and neuroplasticity theoretically linked to chronic SSRI treatments ^53, 67^. They may also provide mechanistic insight into disruptive early side effects (e.g. anxiety) ^28^ and the immediate effects of SSRIs on affective bias that could theoretically contribute to and predict long-term clinical outcomes (e.g. ^41^). Studies of consenting healthy volunteers avoid extraneous confounds of disorders and allow for a powerful within-subject analysis without requiring a placebo control in patients who would otherwise benefit from medication.

It was recognised that the influence of SSRIs could be expressed in different ways across the hierarchical network of interoceptive substrates, which change in function from supporting interoceptive sensory representation (e.g. mid-posterior insular cortex) to their integration into motivational and affective states (e.g. anterior insular cortex and amygdala) ^3^. We, therefore, tested two hypotheses across these networks: First, considering that serotonin tends to generally inhibit exteroceptive processing ^9, 59, 60^, reduce visceral pain ^11, 15, 16^, increase emotional ‘blunting’ ^54, 68^, reduce responses to rewards and punishments ^69^, and reduce amygdala and insula responses to cues of threat (e.g. ^70^) – the primary hypothesis was that an acute SSRI dose would attenuate the general neural response to ordinary internal sensations. The secondary hypothesis was that some SSRI effects on interoception are selective for anxious states. We then undertook a post hoc exploration of the relationship between identified SSRI effects on interoception and acute changes in anxiety.

## Methods and Materials

### Experimental Design

We completed a placebo-controlled, double-blind, randomised cross-over, pharmacological neuroimaging experiment whereby the same participants were tested in two sessions, one following a single dose of an SSRI and one following a single PLACEBO, in random order, separated by a minimum of a seven-day washout period. The SSRI (20mg oral CITALOPRAM (Cipramil, Lundbeck Ltd)) was chosen for its common use, its safety profile, and its exceptionally high selectivity for the serotonin transporter ^71^.

### Participants

Ethical permission was granted by the Brighton and Sussex Medical School Research Governance Ethics Committee (ER/JL332/9). Potential participants were screened with a health questionnaire and a standard psychiatric interview and provided informed consent. Exclusion criteria included: age under 18 years or over 35 years; the presence of significant ongoing medical condition; pregnancy or breastfeeding; currently taking any medication (excluding contraceptive pill); first-degree family history of bipolar disorder; an indication of current or historical mental health disorder, or scanner contraindications (e.g. metallic implants). Participants were instructed to abstain from alcohol or caffeine in the preceding 12 hours before the start of test sessions.

Thirty-one healthy participants were recruited for the study and randomised to a treatment sequence. The sample size was determined by available funding, exceedance of sample sizes used to establish prior acute citalopram effects on neural responses to affective cognition ^41^, our within-subject design, and probable 30% attrition rate. Seven participants withdrew from the study. Neuroimaging data from two participants were excluded due to excessive motion (>6% of volumes identified as motion outliers for either session scan). Neuroimaging data for one participant was excluded due to extreme nausea requiring exiting of the scanner. This left 21 participants measured in each condition (42 sessions total). This group included seven males, had a mean age of 23.9 ± 3.3 (SD), a mean height of 171 ± 9 cm and a mean weight of 64.6 ± 8.9 kg. Before any treatment, these participants experienced a range of trait anxiety levels (State-Trait Anxiety Inventory (trait score) (STAI-T) (mean 34.9 ± 8.3; 11 participants experienced low anxiety (below 35), 9 participants experienced moderate anxiety (between 35 and 44), and one experienced high anxiety (score of 50), and normal interoceptive tendencies (The Body Perception Questionnaire (BPQ), awareness subscale mean 2.4 ± 1.0, stress subscale mean 2.5 ± .88).

### Study Procedure

Participants were tested twice, with separate CITALOPRAM and PLACEBO sessions, at least seven days apart (*M* = 10.3 days, *SD* = 6.87). Assignment to treatment order was double-blind and counterbalanced, with 20mg of oral CITALOPRAM administered in one session and the PLACEBO in another. CITALOPRAM and PLACEBO doses were delivered in identical gelatine capsules filled with microcrystalline cellulose by a pharmacist.

Following the dose, but before testing, participants were given instructions and time to practice the VIA task. To allow CITALOPRAM levels to reach peak absorption, participants had their heartrate recorded with the participant relaxed and sitting. They entered the scanner approximately 3.75 hours after treatment intake. Heart rate variability was measured as the standard deviation of the pulse interval.

State anxiety (STAI-S) was measured before and after the scan session. We used the average to estimate anxiety in the scanner. Similarly, to relate to prior studies and capture change in mood, participant affect was measured with the Positive and Negative Affect Scale (PANAS) before and after scan sessions. The PANAS includes features of state anxiety in addition to other measures of affect. So, only the STAI-S was used to test hypotheses related to anxiety. Participants also completed eight visual analogue scales before and after the scan (the average of these taken for analysis, estimating scanner experiences). Three scales (from 0-100) were given to assess three somatic side effects (nausea, headache and dizziness). Five anxiety-related effects (pairs of antonyms: alert−drowsy, stimulated−sedated, restless−peaceful, irritable-good-humoured, anxious−calm) were used to confirm the STAI-S measures and alert the researchers to excessive side effects.

### Visceral Interoceptive Awareness Task

The visceral interoceptive awareness (VIA) task (Figure 1) has been used previously to identify changes in neural interoceptive processing in multiple clinical contexts ^20, 49, 63–66^. The *Heart* and *Stomach* cue blocks corresponded to interoceptive conditions, and the *Target* blocks to an exteroceptive (visual) baseline condition. In the *Target* condition, after a short interval, the target started changing from black to grey and cycled between shades for a randomly varied time between 0.7s and 1.1s, designed to mimic the approximate frequency of normal heart rate. In half the trials, participants were asked to rate the intensity of the sensations from the area of focus (interoceptive trials) or the intensity of the colour change (exteroceptive trials). These responses helped to ensure that participants focused but were too infrequent for robust statistical inference. The trials selected for intensity rating were randomly re-selected (same for every participant). Sensory condition order (heart, stomach, target) and shades of grey for the TARGET blocks were the same for every participant in a randomised sequence. There were 15 trials in each condition for a total task duration of 15 minutes.

**Figure 1:**
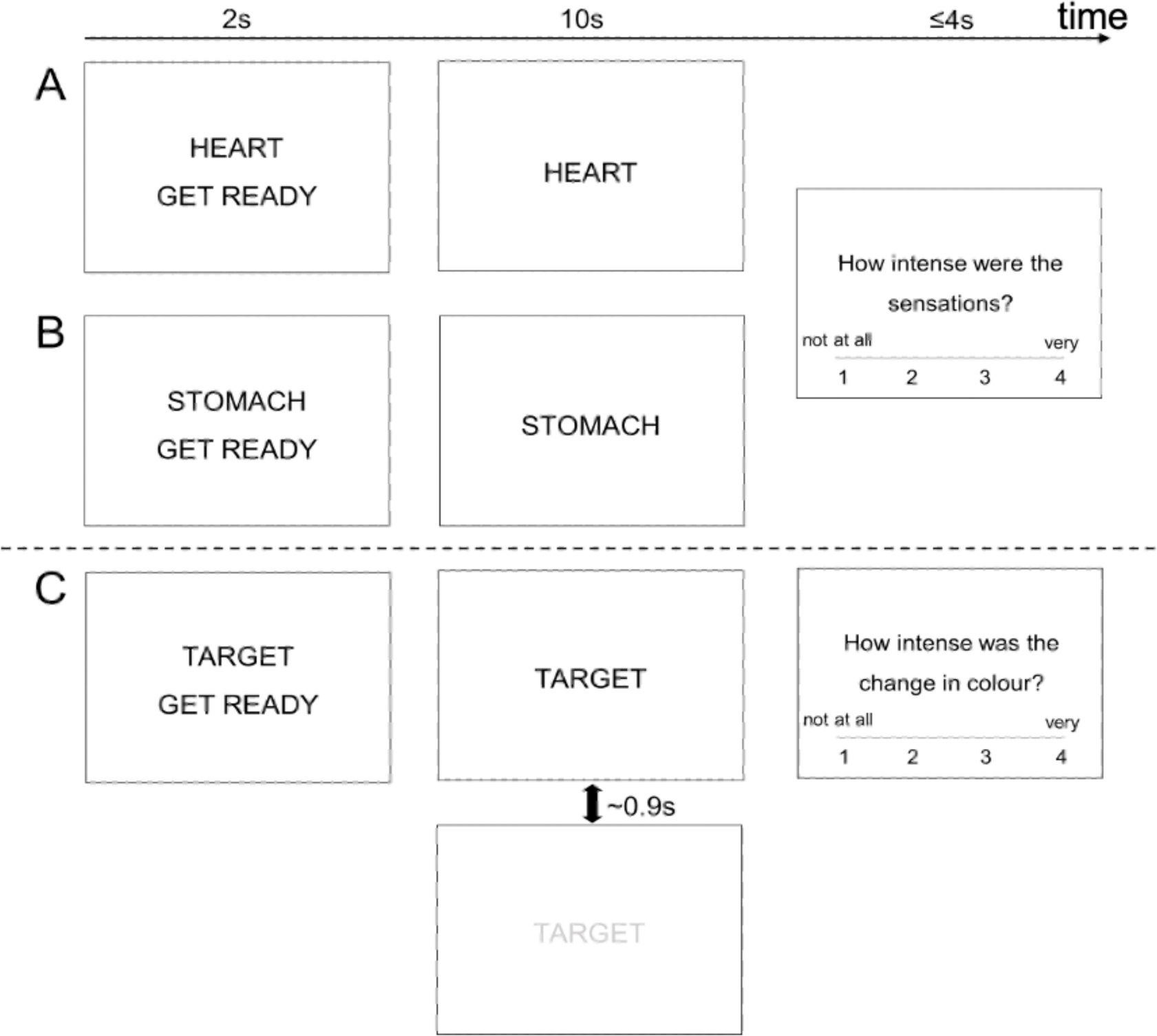
Visceral Interoceptive Attention Task focusing on the (A) heart, (B) stomach, and (C) exteroceptive condition. Each condition was presented 15 times, and the rating phase was presented eight times for conditions A and B and seven times for condition C.

### Neuroimaging Procedures

Magnetic resonance imaging data were acquired using a Siemens Magnetom Prisma 3T scanner (Siemens Healthcare GmbH, Erlangen, Germany) and a 32-channel head coil. We acquired functional magnetic resonance images (fMRI) (T2*, TR=1.5s) during VIA task performance, field maps, perfusion images to test for treatment effects on cerebral blood flow, and T1-weighted structural images for registration. Functional MRI images were pre-processed with a standard pipeline and ICA denoising. See Supplemental Material for further details.

### Image Acquisition

Functional magnetic resonance imaging (fMRI) was acquired using T2*-weighted echo-planar imaging sensitive to blood oxygenation level-dependent (BOLD) signal changes (multiband factor 4, echo time 37ms, repetition time 1500ms, voxel size 2.2 × 2.2 × 2mm, 104 × 104 voxels per slice, 72 slices, field of view (FOV) 205 × 205mm^2^, flip angle 52°), with a varying number of volumes according to the speed of task completion (*M =* 580, *SD* = 36.8). Before the task fMRI sequence on both test sessions, pairs of phase-encode reversed images were acquired for distortion correction (FOV 228 × 228mm^2^ 104 × 104 voxels per slice, echo spacing 0.54ms). In one session, a T1-weighted magnetization-prepared rapid acquisition gradient echo (MPRAGE) structural image of the brain was acquired (echo time 2.2ms, repetition time 2400ms, 0.8mm isotropic voxels, 300 × 320 voxels per slice, 208 slices, FOV 256 × 256mm^2^, flip angle 8°).

### Pre-processing

Functional MRI images were pre-processed with a standard FSL pipeline, including motion correction using the middle volume of the time series as a reference volume, identification of motion outlying volumes using FSL Motion Outliers (root mean squared intensity difference of adjacent volumes method), distortion correction using phase-encode reversed image pairs (using FSL tool top-up), high-pass filtering at 80 seconds, brain extraction, and co-registration to participant structural images. Structural images were co-registered to standard MNI space. Independent Component Analysis (ICA) decomposition was carried out using FSL-MELODIC on unsmoothed data, and denoising was carried out manually, with noise components identified by an independent researcher using published criteria ^72^. Spatial smoothing was completed with a 5mm full-width at half maximum (FWHM) Gaussian kernel.

### Cerebral Blood Flow Change

Changes in resting perfusion that may accompany the pharmacological treatment and confound task-related changes in BOLD signal were measured with pulsed arterial spin labelling (ASL) images acquired using a FAIR-QII sequence (4 label-control image pairs, echo time 16ms, repetition time 4600ms, voxel size 1.5 × 1.5 × 3mm, 126 × 128 voxels per slice, 40 slices, interleaved slices, FOV 192 × 192mm^2^, flip angle 180°, inversion time 1990ms, bolus duration 700ms).

ASL images were processed using Bayesian Inference for Arterial Spin Labelling to produce perfusion-weighted images. ASL images were analysed at the group level in a paired t-test of CITALOPRAM and PLACEBO sessions using Statistical Parametric Mapping (SPM12), with a cluster-wise family-wise error rate of *p* < .05, and with an additional exploratory threshold of *p* < .001 with extent > 50 voxels per cluster.

### Analyses

Analyses of treatment differences in self-reported sensation intensities during the VIA task, heart rate, STAI-S and PANAS scores and VAS scales were carried out using paired t-tests.

Functional images were analysed using FSL FEAT v6.0.3 with two-level generalised linear models (GLM). The first-level models corresponded to individual scan sessions. These included events corresponding to trial conditions (*Heart*, *Stomach* and *Target*) along with regressors of no interest: intensity rating periods, button presses, outlying motion volumes and temporal derivatives of each event. Neural responses to exteroceptive focus (Target) served as high-level controls. They were subtracted from responses to interoceptive focus (Heart, Stomach) to generate contrast images of the relative interoceptive response (IR) to heart focus (heart-IR) and stomach focus (stomach-IR). IRs control for treatment effects on sensory processing, in general.

We completed three second-level analyses. Hypothesis 1 was tested by comparing heart-IR and stomach-IR between treatment conditions. This used FSL’s paired two-group difference design (i.e., a multilevel mixed effect model with a random intercept) with FMRIB’s Local Analysis of Mixed Effects (FLAME 1 & 2) over heart-IR and stomach-IR contrasts of all 42 scan sessions. Fixed factors were treatment (PLACEBO, CITALOPRAM) and test session (1^st^ or 2^nd^). Hypothesis 2 was tested by a model designed to test for anxiety-dependent effects and how the relationship between anxiety and interoceptive response (heart-IR and stomach-IR) changed on CITALOPRAM. This was similar to the first analyses, but adding centred STAI-S scores (which vary between sessions within subjects) and state anxiety-by-treatment interaction (STAI-S scores multiplied by +1 for CITALOPRAM and −1 for PLACEBO) regressors. A drug-induced change in the relationship between anxiety and IRs was tested by one-sample t-tests over the anxiety-by-treatment interaction for both heart-IRs and stomach-IRs of all 42 sessions. To confirm that any significant anxiety-by-treatment interaction effect changed an existing relationship between anxiety and interoceptive response (without CITALOPRAM), replicating prior studies, a third whole-brain search was conducted across all participants in the PLACEBO condition, using regressors of the intercept, test session, and mean-centred STAI-S scores. A one-sample t-test across the latter was used to identify baseline unmedicated relationships between heart-IR and anxiety.

Unless otherwise specified, all reported findings at the second level were whole-brain searches, cluster-corrected using a conservative voxel-wise threshold of Z > 3.1 and a cluster significance level of p <0.05. However, we applied small volume correction where a whole-brain search failed in the main heart-IR comparison. Since we had strong predictions that effects would be observed in or around the amygdala ^50^ and insular cortex ^49^, we generated masks for these regions with the Oxford-Harvard cortical and subcortical probabilistic atlases (using any probability of including the designated region). Only two masks were tested: bilateral amygdalae and bilateral insular cortices (including nearby planum temporale, Heschl’s Gyrus, operculum).

Session order effects were tested independently to establish no confounding effects of the session or session x treatment interaction.

Post-hoc, significant clusters from the treatment effects of the first second-level model were tested for association with increases and decreases of anxiety between sessions after controlling for anxiety-associated side effects and treatment order. Criteria for reporting was either a significance level beyond a Bonferroni corrected threshold (p<0.0083) or a similar effect in the same region on the other side of the brain, which would be very unlikely to occur by chance.

Supplemental tests examined the robustness of effects against side effects of the drugs, session effects, and guess of treatment condition. A separate supplemental regression was also conducted between treatment effects on an independent measure of cardiac interoceptive awareness reported elsewhere^21^ and treatment effects on heart-IR. For robustness tests of all results, please see Supplemental Material (Figures S3-S8, Tables S1-S4).

## Results

### Heart Rate and Subjective Experience

Heart rate was lower on CITALOPRAM, consistent with the effective absorption of CITALOPRAM^73^. There was no change in a specific, subjective experience (Table 1). However, participants rated the likelihood of having received CITALOPRAM higher on a scale of 1-100 when they had received CITALOPRAM (t(20)=2.42, *p =* 0.025), indicating a potential summative influence of subjective effects on conscious awareness of the drug condition. See Supplemental Material for a thorough investigation into the robustness of results relating to this effect.

**Table 1.** fMRI Effect Clusters. Contrast, region (Oxford Harvard Cortical and Subcortical Atlas), number of voxels in cluster, peak voxel z score and peak coordinates (x,y,z) in MNI space.

| **Contrast and Anatomy** | **Voxels** | **Z MAX** | **X** | **Y** | **Z** |
| --- | --- | --- | --- | --- | --- |
| **Citalopram Effect on Stomach-IR** |  |  |  |  |  |
| right posterior insular cluster: planum temporale peak extending to Heschl's gyrus, central operculum and posterior insular cortex | 609 | 4.85 | 44 | -30 | 10 |
| left posterior insular cluster: parietal opercular cortex peak extending to Heschl's gyrus, planum temporale, central operculum and posterior insular cortex | 155 | 4.35 | -38 | -30 | 20 |
| left amygdalar cluster: amygdala peak extending to the anterior insular cortex, parahippocampal gyrus | 135 | 4.35 | -22 | 2 | -18 |
| right amygdalar cluster: frontal orbital cortex peak extending into the amygdala | 107 | 4.23 | 20 | 8 | -16 |
| **Citalopram Effect on Heart-IR (within Amygdala Mask)** |  |  |  |  |  |
| right amygdala, orbitofrontal cortex, parahippocampal gyrus | 67 | 4.28 | 16 | 0 | -22 |
| left amygdala, hippocampus | 23 | 3.74 | -22 | -10 | -18 |
| **Citalopram x State Anxiety Association with Heart-IR** |  |  |  |  |  |
| frontal orbital cortex, insular cortex, inferior frontal gyrus | 341 | 4.53 | 44 | 28 | -8 |
| occipital cortex | 216 | 5.49 | 34 | -88 | -10 |
| frontal orbital cortex, insular cortex | 118 | 4.4 | -38 | 24 | -4 |
| lateral, occipital cortex, precuneus | 109 | 4.31 | 10 | -82 | 48 |
| occipital pole | 86 | 5.22 | -36 | -94 | 4 |

### Cerebral Blood Flow

Analysis of ASL image pairs showed no significant clusters at the familywise error rate, suggesting that CITALOPRAM did not affect cerebral blood flow. Therefore, any effects on BOLD responses were unlikely to be mediated by general effects on blood flow in the brain. With an uncorrected threshold, a single significant cluster was shown in the occipital cortex (peak voxel MNI coordinate: (18, −96, 14), *p*(*uncorrected*) = .001, *pFWE* = .10)

### Interoceptive Attention: Functional Magnetic Resonance Imaging

Relative to PLACEBO, CITALOPRAM reduced the relative interoceptive response to stomach sensation (stomach-IR) within the amygdalae, bilateral posterior insular cortex, and neighbouring regions (Table 1, Figure 2) using a whole brain search. These are referred to henceforth as (left and right) *amygdalar* clusters and *posterior insular* clusters, respectively. During heart focus, treatment differences did not reach the same threshold as the stomach focus when correcting for multiple comparisons across the whole brain. Using small volume correction within the amygdalae and insula cortices, CITALOPRAM was shown to reduce the heart-IR within the right and left amygdalae (Figure 3, Table 1), but there was no evidence for a similar reduction in the insular cortices. A separate analysis demonstrated no order effects within suprathreshold clusters of BOLD activity affected by CITALOPRAM.

**Figure 2.**
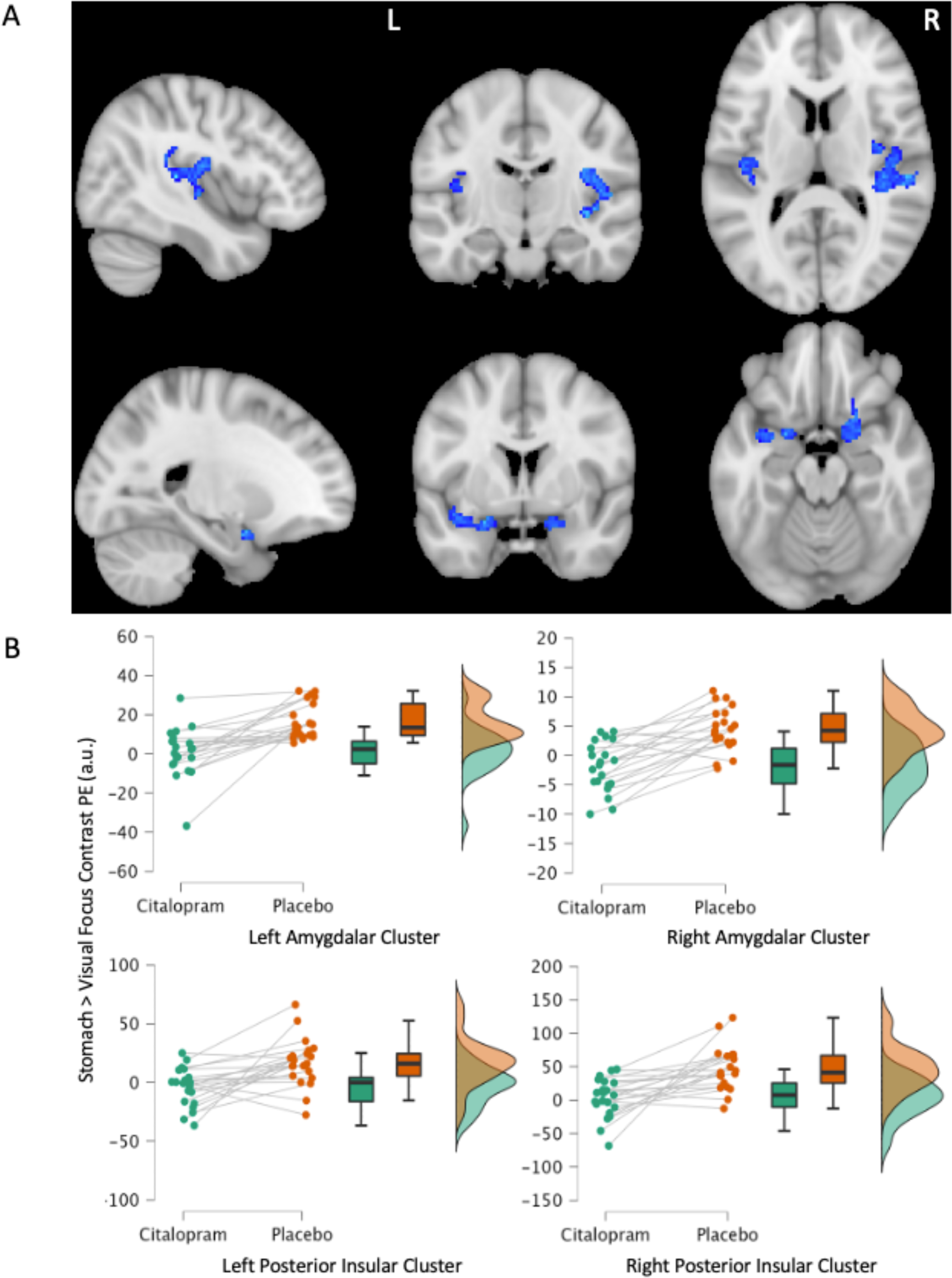
CITALOPRAM Effect on the Interoceptive Response to Stomach Sensation. **A.** Deactivation following 20mg of CITALOPRAM (< PLACEBO) during stomach focus (> visual). Deactivation maps overlayed onto the standard MNI 152 brain (MNI coord (x,y,z): top 43,-15,12; bottom −22, 2, −18) and developed with a voxel threshold of Z > 3.1, and a cluster significance threshold of p < 0.05. Twenty-one participants, repeated measures. Maps generated after controlling for the testing session (1^st^ or 2^nd^). R-L indicate right left. **B.** Plots of average contrast parameter estimates (PE) for each significant cluster. Green= CITALOPRAM. Orange = PLACEBO. a.u. = arbitrary units.

**Figure 3.**
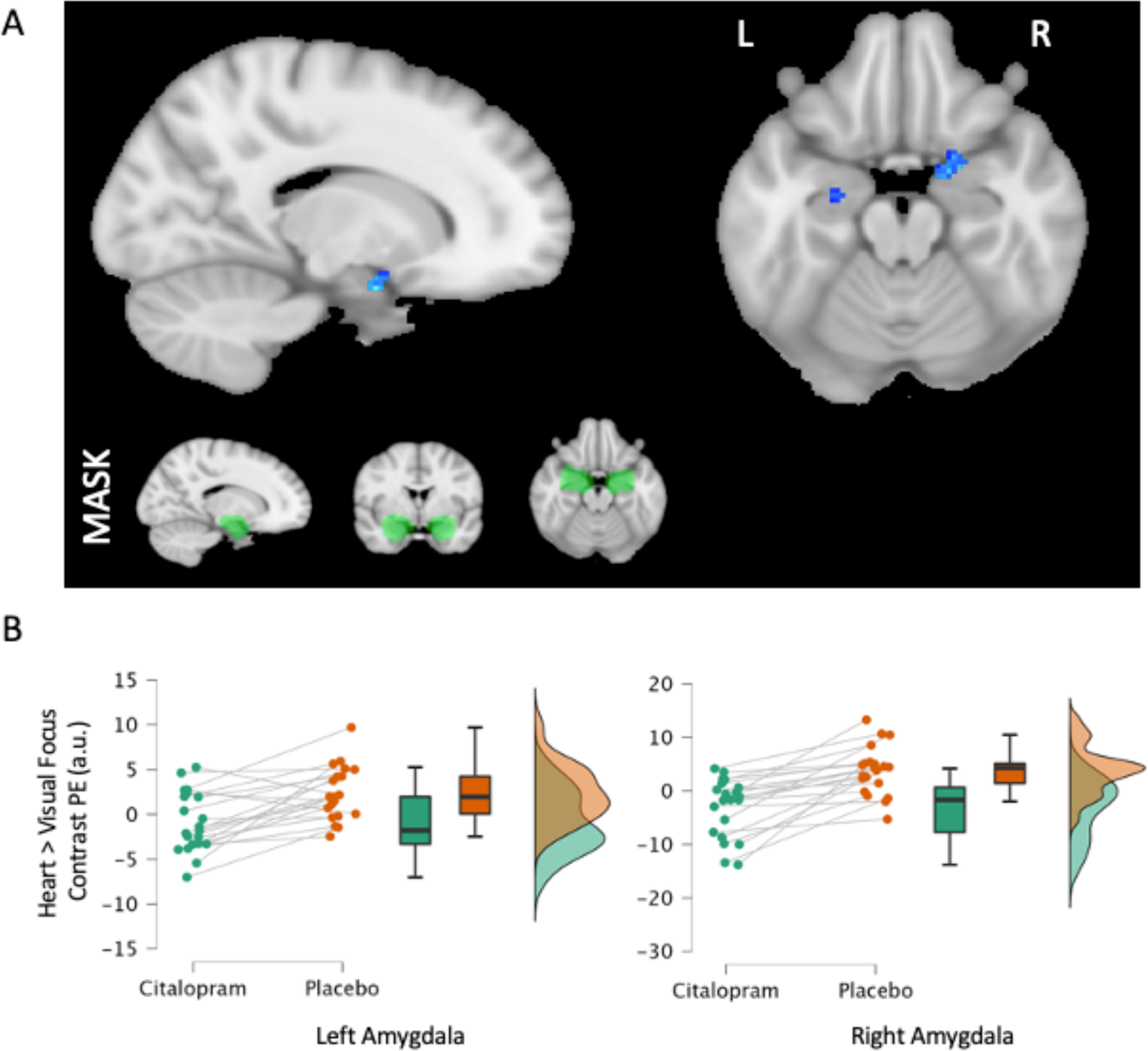
CITALOPRAM Effect on Interoceptive Response to Heart Sensation. **A**. Reduced amygdala activation following 20mg of CITALOPRAM (vs. PLACEBO) while attending to the heart (vs. visual stimuli). Deactivation maps (blue) overlayed onto the standard MNI 152 brain (MNI x = 16, z = −18), developed by a voxel threshold of Z>3.1, a cluster significance threshold of p<0.05, and small volume correction within an anatomical mask of amygdalae with any probability of amygdalae using the Harvard-Oxford Subcortical Structural Atlas (green). **B.** Difference plots of average contrast parameter estimates (PE) (heart-IR) in each drug condition for each significant cluster. a.u. = arbitrary units.

### Anxiety

State anxiety decreased for some participants and increased for others following an acute dose of CITALOPRAM. This resulted in no average anxiety change (Table 1).

In a whole-brain search, we discovered a reduced association between anxiety and interoceptive response for the heart-IR on CITALOPRAM within symmetrical bilateral clusters that spanned anterior insular and posterior orbitofrontal cortices (Figure 4A, Table 1). A visual appraisal of the data indicated that the heart-IR within these regions increased with state anxiety when participants were on PLACEBO, but this relationship was reversed by CITALOPRAM (Figure 4B). In an independent whole brain cluster-corrected analysis of the PLACEBO condition, state anxiety was shown to vary with response to heart-IR in the right anterior insular/orbitofrontal cortex (316 voxels, Peak Z = 5.13, MNI coord: 44,30,-12). On the left, this effect was also present but did not survive cluster correction (58 voxels, p<.001 uncorrected, peak MNI coord −44, 20, 8) (Figure 4A). Further associations between state anxiety and heart-IR were observed on PLACEBO, but only the anterior insular/orbitofrontal clusters demonstrated a reduced association on CITALOPRAM. A similar association was not observed for the stomach-IR.

**Figure 4:**
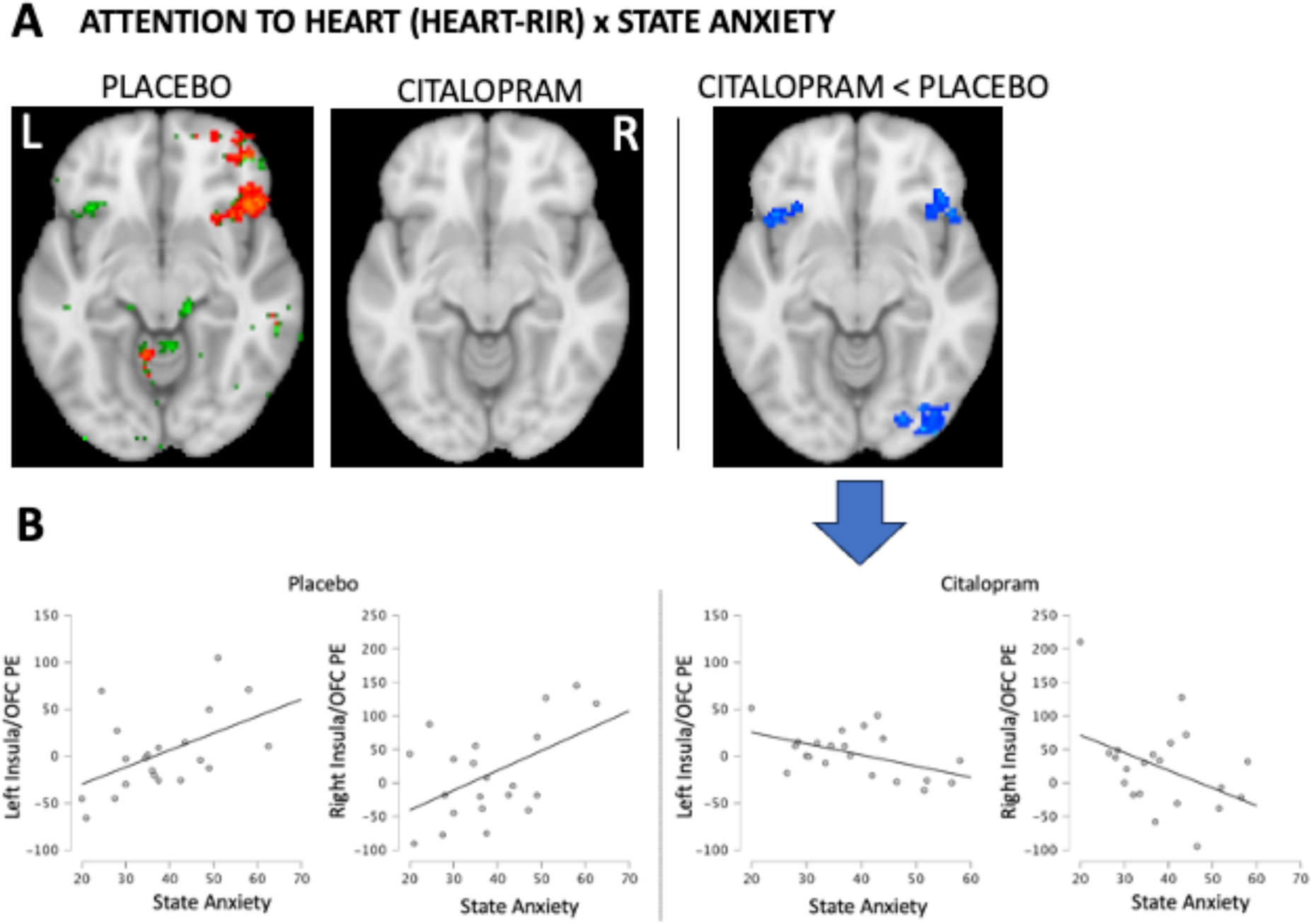
Changing the Anxiety-Interoception Relationship. **A**. Relationship between neural response to heart focus (heart-IR) and state anxiety in PLACEBO and CITALOPRAM conditions, and the significant reduction of relationship CITALOPRAM. Maps overlayed onto the standard MNI 152 brain (MNI coord (z = −10). Red and Blue maps result from whole-brain searches that were cluster corrected, developed by a voxel threshold of Z>3.1 and a cluster significance threshold of p<0.05. Green is from a whole brain search, uncorrected at p<0.001. **B:** Plots of state anxiety (STAI-S) variation with heart-IR average parameter estimates (PE, in arbitrary units) within the frontal orbital cortex / insular clusters of the CITALOPRAM > PLACEBO contrast. Illustration purposes only. Refer to Table 1 for statistical inference.

The state anxiety measure captured anxiety during the scan, which medication side effects could influence. The state anxiety-by-treatment interaction was robust to covariates of nausea, dizziness, headache, guess of drug condition, and the interactions of these measures with the treatment condition. To further rule out the influence of side effects, we tested and found that trait anxiety (STAI-T), measured before any treatment, also predicted the effect of CITALOPRAM on heart-IR within the same bilateral orbitofrontal/insular cortex clusters (Table S1, Figure S2). This supports the conclusion that variation in heart-IR has a source in individual differences in anxiety.

Changes in state anxiety were associated with changes in nausea (r = .68, p <.001), headache (r = 0.54, *p =* 0.014), and dizziness (r = .45, *p =* 0.047) after controlling for treatment order. In the post-hoc examination of the main effects, the remaining variance in state anxiety change after controlling for these associations was associated with changes in stomach-IR within the amygdalae (average extracted parameter estimates left amygdala cluster b = 2.2, t(15) = 3.1, *p =* .008, r = .52). A conservative Bonferroni corrected alpha level for this post hoc analysis (6 tests) is p = .0083. The right amygdala effect (b = .5, t(15) = 2.2, *p =* .044, r = .49) is reported due to the same effect on the left. See Figure 5.

**Figure 5.**
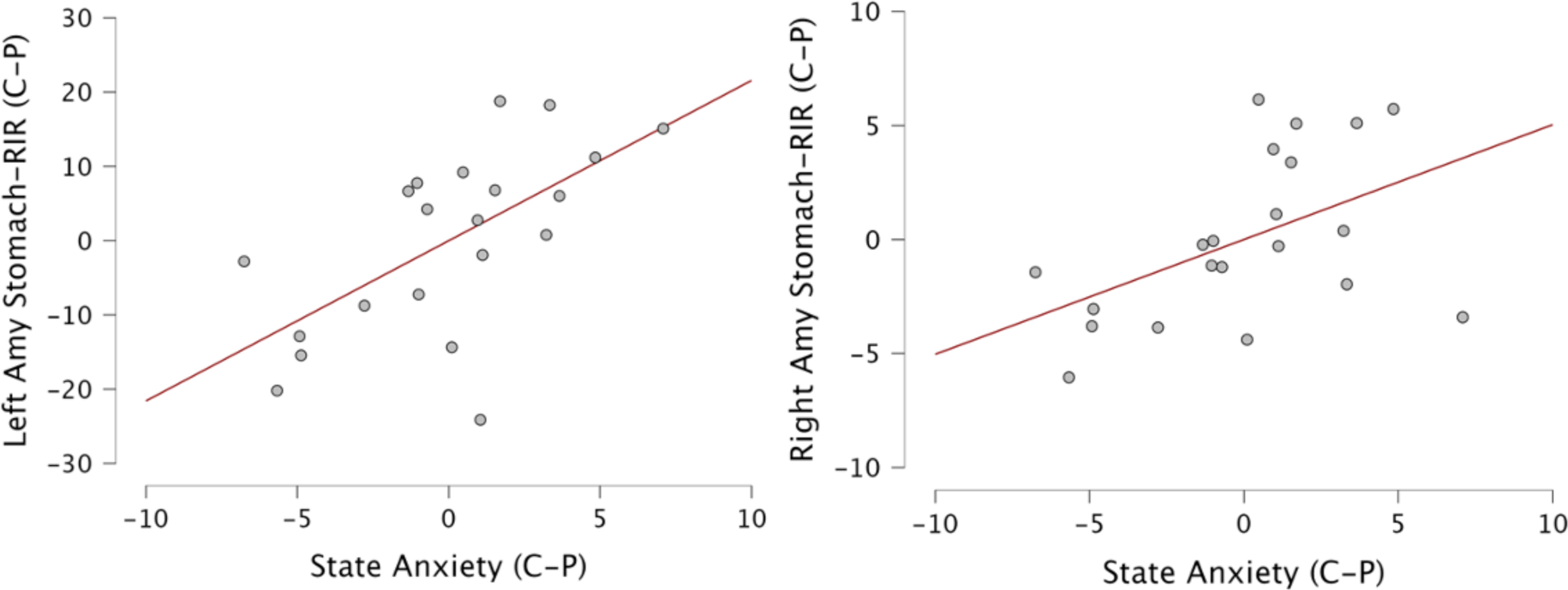
Association Between CITALOPRAM’s Effect on State Anxiety and Its Effect on Stomach-IR. These partial residual plots show the change in state anxiety and change of stomach-IR (extracted average contrast parameter estimates in stomach-IR amygdalar clusters (Table 1), with effects of nausea, headache, dizziness, and treatment order also in the model. C-P is CITALOPRAM – PLACEBO. Right Amy = amygdalar cluster.

### Interoceptive Insight

In a previous study involving contributions from the same participants, a single CITALOPRAM dose increased conscious awareness of the reliability of inferences made from cardiac interoception ^21^. We explored whether this effect correlated with the SSRI-induced reduction of heart-IR, which it did within the left (but not right) amygdala (Supplemental Material, Figure S1).

## Discussion

A single dose of the SSRI CITALOPRAM reduced the relative neural response to attended normal internal sensation in viscerosensory (e.g. posterior insular cortex) and integrative/emotion-processing regions (e.g. amygdala) of interoceptive processing pathways ^3,74^. The posterior insular cortex, in which CITALOPRAM reduced stomach-IRs, is distinct from mid or anterior insular regions in its connectivity and cytoarchitecture ^3,75^. It is the predominant destination for topographically organised viscerosensory information from the thalamus, originating from ascending sympathetic and parasympathetic pathways from body tissues ^1,48, 75^. Neuroimaging studies demonstrate reliable activation of this insular region by gastric sensation ^34, 76^, suggesting that increased extracellular serotonin reduced primary sensory processing of the upper gastrointestinal tract. Theoretically, this basic sensory information then travels through hierarchical networks to the mid and anterior regions of the insular cortex, where it is integrated into conceptual interoceptive representations through interaction with the prefrontal cortex, amygdala and striatum to influence allostasis, motivation, emotion, and related cognitive processes ^3, 48, 74, 77^.

In the amygdalae, both stomach-IRs and heart-IRs were reduced by CITALOPRAM. The amygdalae are innervated by serotonin projections from the raphe nuclei and receive interoceptive information via ascending viscerosensory pathways of the brainstem, the thalamus, and interconnections with the cortex (strongly connecting with anterior insula cortex) ^75, 76, 78, 79^. Here, the interoceptive information is proposed to steer allostasis by influencing arousal and salience attribution ^17, 74, 77, 80^. In the present study, the reduction of IRs could reflect reduced interoceptive input to the amygdala or a change in how the amygdala uses interoceptive sensations.

Prior research demonstrated that individuals with greater anxiety exhibit a greater right ^43^ and left ^44^ anterior insula cortex response to attended heart sensation. In the present study, we confirmed this state-dependent relationship in the PLACEBO condition in the anterior insular cortex (overlapping on the left, but ventral to previous findings on the right). The effect of CITALOPRAM, correspondingly, was also state-dependent. Heart-IR in the anterior insular and adjoining orbitofrontal cortex was reduced in proportion to anxiety levels, breaking the anxiety-interoception relationship in this region. This effect represents a reduction in the link between anxiety and interoception, not an anxiety reduction. Reduced links between anxiety and behavioural measures of interoception have been previously associated with increased anhedonia ^56^. One might, therefore, wish to explore whether SSRI-induced changes in the anxiety-interoception relationship in the anterior insula relate to emotional blunting associated with SSRI treatment in some individuals ^54^. Moreover, anxious states have been proposed to relate to accumulating relationships between levels of anxiety, heightened interoceptive responses, and further anxiety about those heightened interoceptive responses ^30, 41^. Therefore, this finding sets the stage for investigations of whether serotonergic perturbation of the anxiety-interoception relationship could have a role in a variety of long-term SSRI treatment outcomes. More generally, however, this result confirms that some modulatory effects of serotonin on interoceptive processing are state-dependent, as they are suggested to be for exteroceptive processes.

As expected from prior research, anxiety responses increased for some and decreased for others, resulting in no net change ^28, 41, 50^. However, we have now provided preliminary insight into the individual differences in response, suggesting that the amygdala’s response to stomach sensation may relate to increases and decreases in anxiety following an SSRI dose. The bilaterality of the effect provides confidence in this result, so if replicated in a larger sample, this could underscore the importance of considering links between anxiety and gastrointestinal sensation in psychiatric and internal medicine research ^32, 81, 82^.

Like similar experimental medicine studies, we employed a single-dose SSRI protocol to better understand the cognitive impact of acute increased endogenous serotonin ^41, 50^. Acute effects of SSRIs can differ from effects after seven or more days due to desensitisation of autoreceptors and other adaptive effects ^67, 83^. Additional research would therefore be needed to extrapolate to the longer-term effects of SSRI treatment. Secondly, this was a study of young, healthy volunteers. This provides controlled inferences about normal function unencumbered by interactions with symptoms or other medications and avoids ethical challenges of within-subject pharmacological study in a patient group. However, only with further research should one extrapolate these findings to clinical contexts and experiences across the lifespan. There is also much more to learn about the precise mechanisms of these effects. Citalopram has exceptional selectivity for the serotonin transporter. However, the role of knock-on effects on other neurotransmitter systems and the roles of specific serotonin receptors remains unknown.

Overall, we found that an acute increase in extracellular serotonin reduces central neural responses to interoceptive information in young, healthy individuals in both general and anxiety-dependent fashion. This opens new avenues of research in other populations and contexts for a better understanding of interoception and its relationship to affective states.

## Data Availability

All data produced in the present study are available upon reasonable request to the authors. Summary data is available at DOI 10.6084/m9.figshare.22786274.

https://figshare.com/s/88d2088d017407a60574

## Acknowledgements

We would like to thank Dr. Jason Avery for his advice on the VIA task at the early stages of this project. We would also like to thank Dr. Andrew Barritt and Dr. Iulia Bogdan for providing medical cover. This study was sponsored and funded by the University of Sussex.

## Author contributions

Conceptualization: DCM, JJAL

Methodology: DCM, JJAL, HC, SG

Investigation: JJAL, KA

Data preparation: DCM, JJAL, LS

Statistical Analysis: DCM, JJAL

Project administration: JJAL, DCM

Supervision: DCM

Writing – original draft: DCM, JJAL

Writing – review & editing: DCM, JJAL, LS, HC, SG, LS

