## Supplemental Material for "General and Anxiety-Linked Influence of Acute Serotonin Reuptake Inhibition on Neural Responses Associated with Attended Visceral Sensation"

**Affiliations:**

All work was completed at the University of Sussex.

**Instructions to Participants for Interoceptive Attention Task.**

Before the experimental trials, participants were given the following instructions:

'*While the word 'HEART' or 'STOMACH' is shown on the screen, focus attention on the intensity of the sensations experienced from the area of your heart or stomach. When the word 'TARGET' is shown on the screen, it will sometimes change from black to different shades of grey. Focus attention on the amount that the colour changes. When a CROSS is shown on the screen, you can rest. Please keep your eyes open and try not to think of anything in particular. You will sometimes be asked to rate how intense the sensations or the colour changes were, using the buttons shown. Please make sure that you breathe evenly throughout the task. You must NOT hold your breath at any time.'*

**Relationships Between Clusters**

Across participants, average posterior insula cluster responses to CITALOPRAM while focusing on the stomach, on the left and right, were highly correlated, sharing 84% of their variance (r = .92, p < 0.001). Amygdala clusters were less correlated with each other (r = 0.57, $p$ = 0.007), and with insula responses (left amygdala cluster / left posterior insula cluster: r = .56, r = .47, $p$ = 0.03; right amygdala cluster / right post insula cluster: r = .58, $p$ = 0.006), each sharing around 32% of variance. Similarly, during heart focus, amygdala cluster changes shared only around 28% of variance (r = .53, $p$ = .014).

**Cardiac Interoceptive Insight**

Participants also performed an independent heartbeat discrimination task outside the scanner (for a full description of this task and protocol see (1), to which the present data contributed). During each of 20 trials (performance on 20 trials correlate at r = .7 with performance on 100 trials (2)), their heartbeat was measured in real-time, while a computer played a set of ten tones at either 250 ms or 550 ms after the R-wave corresponding to judgements of maximum and minimum simultaneity. The participant was directed to respond to whether the tones were in or out of time with their heartbeats and how confident they were in that answer using a VAS scale ranging from 'total guess' to 'complete confidence' on a scale of 1 to 10. Metacognitive cardiac interoceptive insight is measured as the ability of confidence to classify correct and incorrect responses. In a larger sample to which this project contributed, CITALOPRAM increased this ability. One participant of the present dataset was removed due to feeling their pulse in the finger. CITALOPRAM's effect on cardiac interoceptive insight was specifically tested for association with neural effects during heart focus, to bridge behavioural and neural effects. Change of activity within the left, but not right, amygdalar cluster on CITALOPRAM was associated with increased interoceptive insight (r = -.48, *p* = 0.033). This effect remains after controlling for change of heart rate (r = -.55, *p* = .016) or treatment order (r = -.47, *p* = 0.039), *p* = .021).

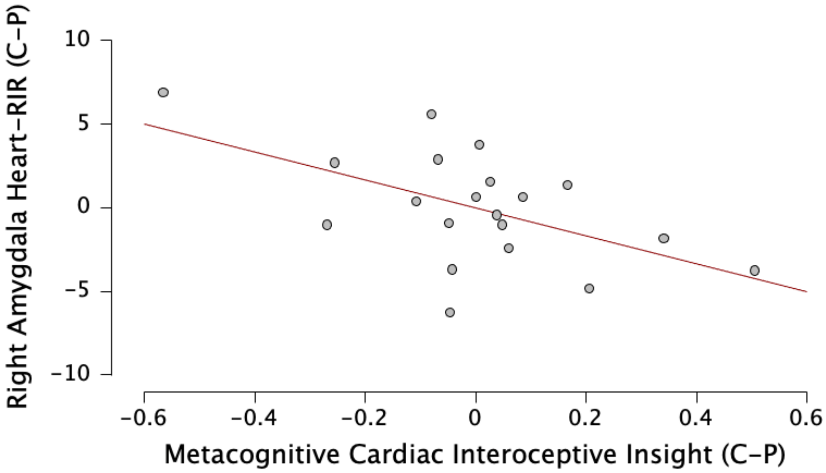

**Figure S1. Change in Metacognitive Cardiac Interoceptive Insight and Neural Response to Heart Focus in the Left Amygdala (N=20)**. C = CITALOPRAM, *P* = PLACEBO.

### Robustness Tests

#### CITALOPRAM's Interaction with State and Trait Anxiety

The interaction effect of CITALOPRAM and state anxiety to generate an increased *reduction* of heart-RIR in the anterior insular/orbitofrontal cortex is unlikely to be caused by side effects because increased visceral sensation would be expected to increase the neural response in these regions. Only an association with dizziness was found (dizziness r = .38, $p$ = .015; nausea r = .251, $p$ = .118; headache r = .226, $p$ = .160) and only if uncorrected for multiple comparisons). However, one may also avoid attending sensations if feeling side effects. This could result in a reduction of heart-RIR. To test for independence of these side effects, we constructed linear mixed models of their heart-RIR using extracted parameter estimates from significant orbitofrontal/anterior insular clusters (Figure 3). These models included random intercepts and fixed effects of treatment (PLACEBO/CITALOPRAM), session, STAI-S scores, treatment x STAI-S interaction, and main effects of nausea, headache, dizziness and each of their interactions with treatment. For both clusters, the STAI-S x treatment interaction remained strongly significant (*ps* < .001), despite covariates.

While STAI-S best captures anxiety levels at that moment, the trait anxiety measure, STAI-T, captures how one *generally* feels. STAI-T scores were measured before any treatment was received and therefore unaffected by the side effects of the drug. STAI-T scores were accordingly not related to headache, nausea, or dizziness (*p*s > .3). If the interaction effect of STAI-T and treatment overlaps with the STAI-S x treatment interaction effect on heart-RIR, then this would provide further assurance that the latter is related to anxiety that is independent of the SSRI's side effects.

A linear mixed model (random intercept, fixed effects of treatment, STAI-S, their interaction, and treatment order) demonstrated that STAI-T scores similarly moderated the effect of CITALOPRAM in the right (b = -3.39, t( 21) = -4.4, p < .001) and left ( b = -1.7, t(21) = -3.1, $p$ = .003) frontal orbital cortex / insular cortex clusters of Figure 3. These effects are unaffected by the addition of nausea, headache, dizziness and/or their interaction with CITALOPRAM as additional fixed effects, and these latter effects were not significant.

For further assurance, the interaction of trait anxiety (STAI-T) and CITALOPRAM was investigated as a parametric predictor of CITALOPRAM effects, with a three-level analysis in FSL. Lower-level analyses were contrasted within each subject at a second level to generate contrast images for each participant that represented differences in activity between treatment conditions. These contrast images were then elevated to a third-level analysis, which allowed for the test of association of differences with variation in STAI-T scores, entered as a mean-centred covariate, together with a variable of treatment order. In a whole brain analysis, STAI-T scores predicted drug effects in the same right anterior insular/orbitofrontal cortex cluster on the right as STAI-S. If limiting statistical cluster correction to the regions associated with state anxiety, the effect is also present in the left cluster.

**Table S1. Trait Anxiety (STAI-T) x CITALOPRAM Effect on Heart-RIR.** Harvard-Oxford Atlas label, cluster size, Z score of peak voxels, and MNI coordinates.

| | Voxels | Z MAX | X | Y | Z |
|---|---|---|---|---|---|
| ***Whole brain search:*** | | | | | |
| *Frontal Orbital Cortex / Insular Cortex* | 186 | 4.29 | 48 | 20 | -10 |
| *Frontal Pole* | 157 | 4.29 | 20 | 52 | 22 |
| ***Small volume correction, within STAI-S x DRUG effect clusters:*** | | | | | |
| *Frontal Orbital Cortex / Insular Cortex* | 119 | 4.29 | 48 | 20 | -10 |
| *Frontal Orbital Cortex / Insular Cortex* | 59 | 4.27 | -40 | 18 | -10 |
| *Inferior Frontal Gyrus* | 22 | 4.05 | 52 | 14 | 8 |
| *Inferior Frontal Gyrus* | 18 | 3.81 | 48 | 28 | 6 |
| *Precuneus* | 7 | 3.92 | 6 | -82 | 50 |
| *Lateral Occipital Cortex* | 5 | 3.55 | -26 | -82 | -2 |

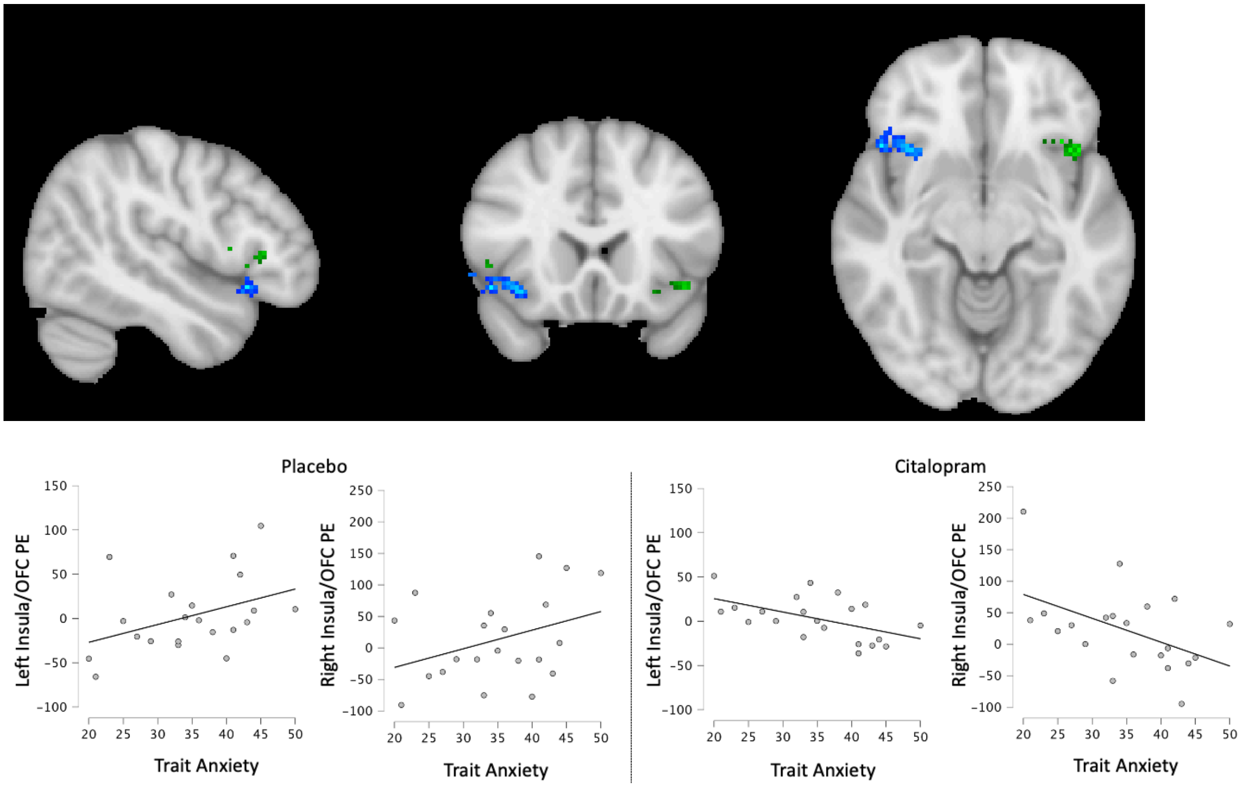

***Figure S2. Effect of CITALOPRAM on Trait Anxiety-Interoception Relationship*: Top:** Reduction of relative neural interoceptive response to heart sensation following 20mg of CITALOPRAM (< PLACEBO) in proportion to trait anxiety (STAIT-T), measured before any treatment. Blue maps are whole brain cluster corrected. The green map resulted from small volume correction within clusters of equivalent contrast using state anxiety (Figure 3). Maps are developed by a voxel threshold of Z > 3.1 and a cluster significance threshold of $p<0.05$. N= 21. **Bottom:** Scatter plots of STAI-T scores to heart-RIR responses in CITALOPRAM and PLACEBO conditions, within frontal orbital cortex / insular clusters generated by the STAI-S x treatment interaction (Table 4). Compare to Figure 3, main text.

### Considering Heartrate

Given that CITALOPRAM reduced heart rate, it was important to understand whether this influenced our results. We ran a *post hoc* repeated measures mediation analysis to test whether the effect of CITALOPRAM on each of the reported responses (extracted average contrast parameter estimates from Tables 2 and 3) is independent of heart rate changes. This includes a test of the drug effect in the presence of a covariate of 'change in heart rate'. The direct effect of CITALOPRAM on neural responses was preserved in all cases ($p < 0.001$); there was no interaction of drug effects with the heart rate change across participants and no mediation by the heart rate change.

**Considering Session Order**

All mixed-effect model imaging results include a covariate for the effect of whether it was the participant's first or second time performing the interoception task in the scanner. In addition, a separate analysis of order effects did not demonstrate any significant effect overlapping with the effects of CITALOPRAM.

However, after participant withdrawals, the sample was not balanced between randomised treatment order conditions: thirteen received the CITALOPRAM in the first session, and eight participants received a PLACEBO in the first session. We sought certainty that this did not influence inferences made from our results. We, therefore, addressed this with a robust dataset with a perfect balance of treatment order, in which all results were preserved (see below).

**Considering Guess of the Treatment Condition**

To indirectly capture the somatic effects of CITALOPRAM and test the preservation of the blind, we asked participants to rate their belief that they had received CITALOPRAM in each session at the time of testing. No participant correctly responded with a certainty of being on CITALOPRAM or PLACEBO. However, a paired t-test demonstrated that the rated probability of being on CITALOPRAM was higher when participants had received CITALOPRAM ($t(20) = 2.42$, $p = 0.025$). This is an imperfect measure because the second session ratings result from different information than the first. Higher difference between CITALOPRAM and PLACEBO condition ratings of being on the drug are higher if CITALOPRAM is received first ($t(19) = 2.3$, $p = .035$). Since the experimenter was blind to the conditions, the increased rated probability of being on CITALOPRAM after receiving it was unlikely due to implicit cues from the experimenter. Regardless, this effect merited further investigation in the robust dataset below.

**Robust Dataset Results**

To address the potential effects of treatment order and insight into drug conditions, we developed a supplementary robust dataset. This group was created by removing participants in the larger treatment order group with the most significant difference of belief of having received CITALOPRAM between drug conditions until the groups were balanced for order (8 participants in each order) and guess of treatment (t(15) = 0.86, $p$ = .40), with a total of 32 test sessions. In this dataset, there was no difference in any subjective measure between treatment conditions ($ps > .25$). Near identical results were found in the robust dataset as found for the full dataset (Figures S4, S5, S6 & S7, Tables S1 & S2 & S3). The only effect losing significance, but remaining on trend, was the association of anxiety change to the right amygdala cluster response to CITALOPRAM (left amygdala cluster effect remains). In general, fMRI effects could, therefore, not be attributed to treatment order or belief in drug conditions.

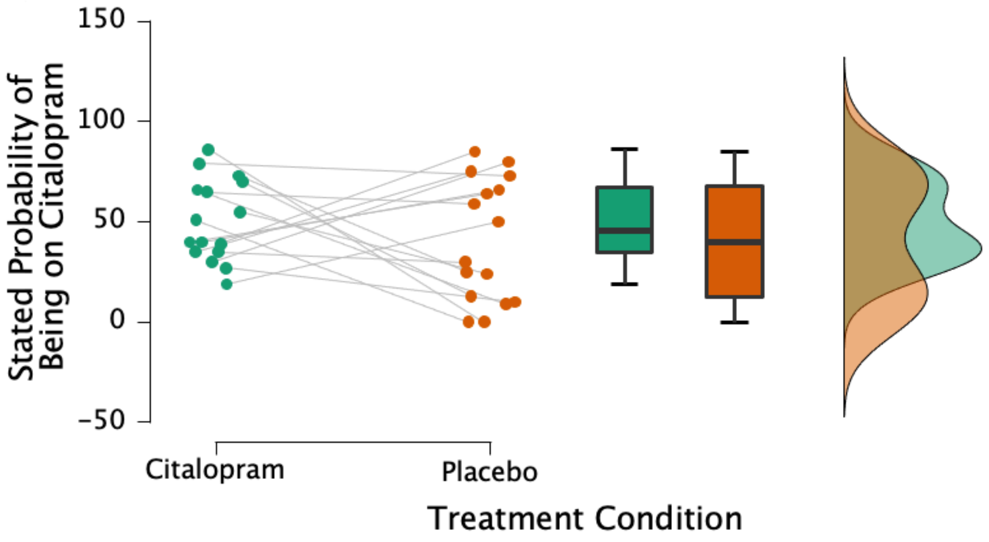

**Figure S3. Stated Probability of Drug Condition, Robust Dataset**

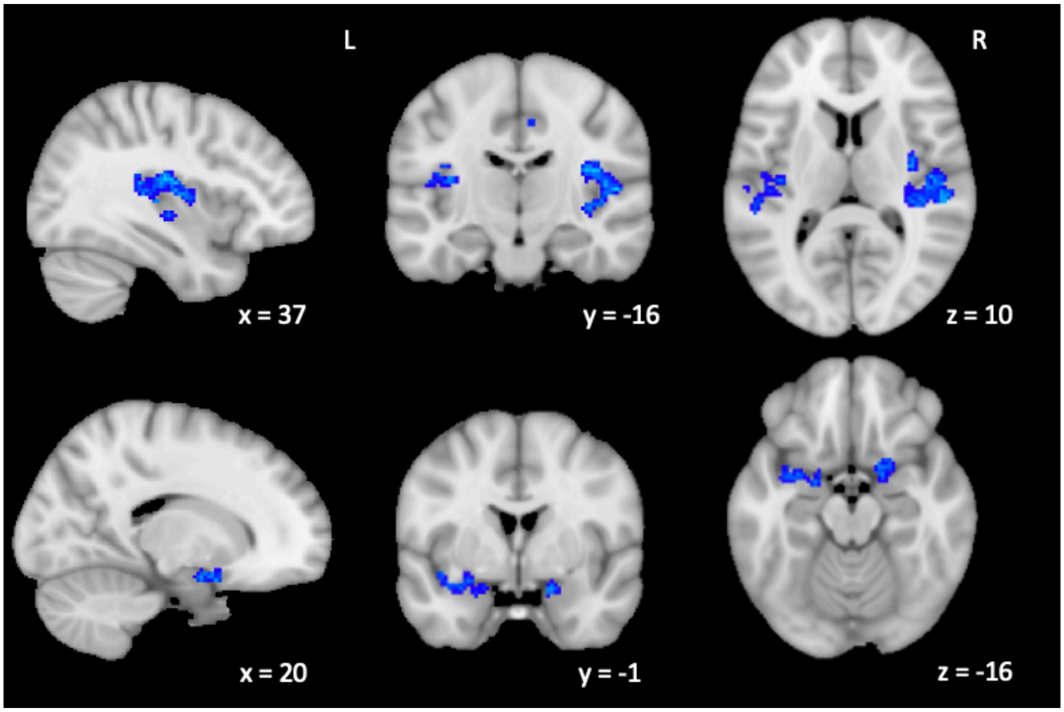

**Figure S4. CITALOPRAM Effect on Stomach Focus, Robust Dataset.** Reduced activation following 20mg of CITALOPRAM (compared to PLACEBO) while attending to the stomach (relative to attending to visual stimuli) is found in an abridged dataset, with 16 volunteers that were balanced for treatment order and self-rated probability of being on the drug between PLACEBO and CITALOPRAM conditions. Clusters are qualitatively the same as in the full 21 volunteer sample. Deactivation maps overlayed onto the standard MNI 152 brain and developed with a voxel threshold of Z > 3.1, and a cluster significance threshold of $p < 0.05$. Maps generated after controlling for drug/PLACEBO session order. R = Right, L = Left.

**Table S2. CITALOPRAM Effect on Stomach Focus, Robust Dataset.** Harvard-Oxford Atlas label, cluster size, Z score of peak voxels, and MNI coordinates.

| REGION | VOXELS | Z MAX | X | Y | Z |
|---|---|---|---|---|---|
| *Right posterior insula cluster: right planum temporale peak, extending to Heschl's gyrus, posterior insular cortex, central opercular cortex* | *742* | *5.13* | *40* | *-34* | *14* |
| *Left posterior insula cluster: left posterior insular cortex peak extending to Heschl's gyrus, insular cortex, central opercular cortex and planum temporale* | *385* | *4.71* | *-36* | *-18* | *14* |
| *precentral gyrus* | *303* | *4.62* | *10* | *-26* | *46* |
| *Left amygdala cluster: frontal orbital cortex peak extending to left amygdala and anterior insular cortex* | *184* | *4.75* | *-24* | *8* | *-22* |
| *cerebellum* | *153* | *4.87* | *-8* | *-76* | *-40* |
| *cerebellum* | *123* | *4.34* | *-10* | *-50* | *-54* |
| *Right amygdala cluster: right frontal orbital cortex extending to right amygdala* | *105* | *4.43* | *20* | *6* | *-20* |

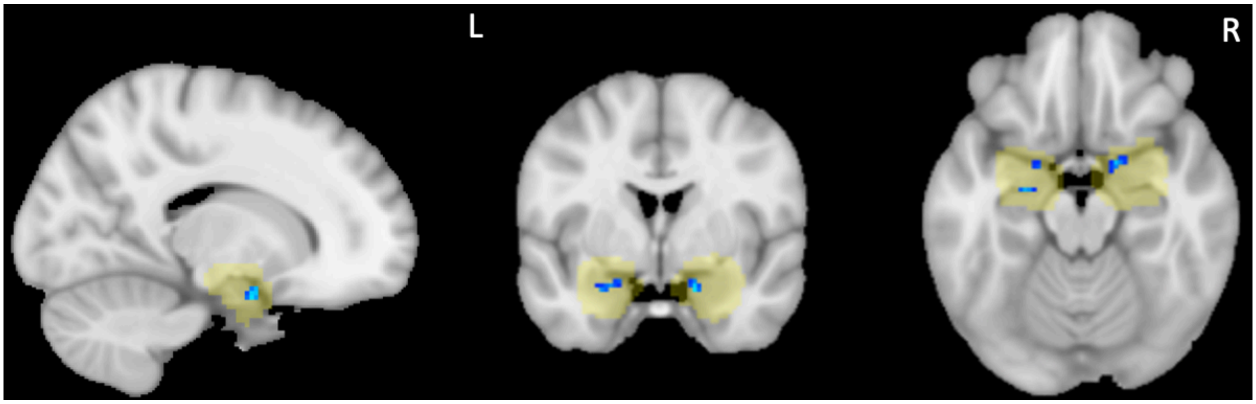

**Figure S5. CITALOPRAM Effect on Heart Focus, Robust Dataset. A.** Reduced activation in the amygdala following 20mg of CITALOPRAM (vs. PLACEBO) while attending to the heart (vs. visual stimuli), with 16 volunteers that balanced for treatment order (8 of each) and self-rated probability of being on the drug between PLACEBO and CITALOPRAM conditions. Deactivation maps overlayed onto the MNI 152 brain (MNI coord (x,y,x): 16,0,-18) and anatomical mask (yellow). R-L indicate right and left.

**Table S3. CITALOPRAM Effect on Heart Focus, Robust Dataset.** Harvard-Oxford Atlas label, cluster size, Z score of peak voxels, and MNI coordinates.

| **Region** | **Voxels** | **Z MAX** | **X** | **Y** | **Z** |
|---|---|---|---|---|---|
| *right amygdala* | 38 | 4.02 | 16 | 0 | -22 |
| | 30 | 4.15 | 24 | -10 | -22 |
| *left amygdala* | 27 | 3.77 | -24 | -10 | -18 |
| | 19 | 3.75 | -24 | 2 | -22 |

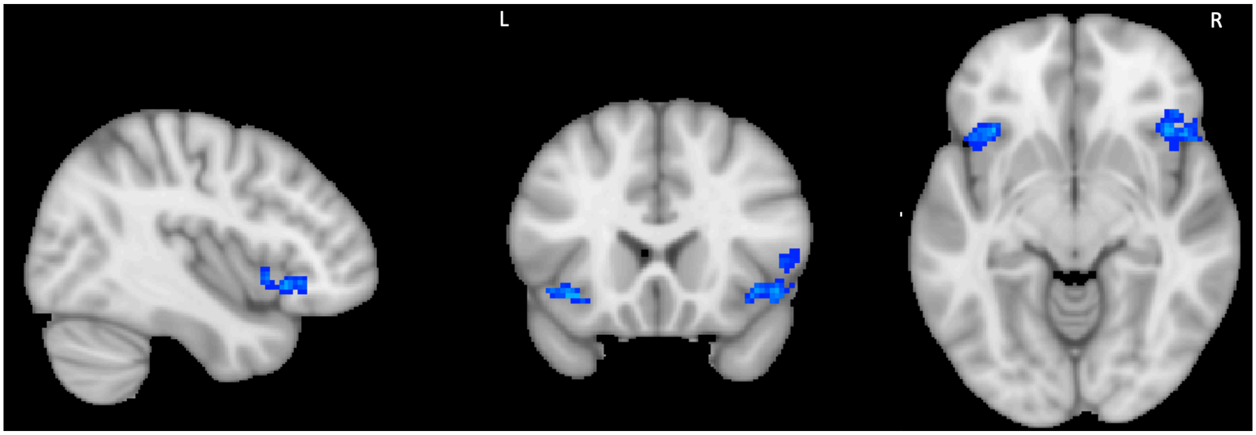

**Figure S6. CITALOPRAM Effect on Stomach Focus x State Anxiety, Robust Dataset.** Reduced activation following 20mg of CITALOPRAM (compared to PLACEBO), while attending to the heart (relative to visual stimuli), in proportion to individual differences in state anxiety, with 16 volunteers that balanced for treatment order (8 of each) and self-rated probability of being on the drug between PLACEBO and CITALOPRAM conditions. Deactivation maps overlayed onto the MNI 152 brain (MNI coordinates (x,y,x): 41,20,-8) and developed by a voxel threshold of Z>3.1, and a cluster significance threshold of p<0.05.

**Table S4. CITALOPRAM Effect on Stomach Focus x State Anxiety, Robust Dataset.** Harvard-Oxford Atlas label, cluster size, Z score of peak voxels, and MNI coordinates.

| Region | Voxels | Z MAX | X | Y | Z |
|---|---|---|---|---|---|
| *frontal pole, right* | 296 | 4.97 | 28 | 58 | 24 |
| *frontal orbitofrontal cortex / insular cortex, right* | 261 | 4.42 | 40 | 22 | -8 |
| *lateral occipital cortex, superior division, right* | 143 | 4.61 | 50 | -70 | 32 |
| *frontal pole, left* | 121 | 4.61 | -20 | 52 | 36 |
| *frontal orbitofrontal cortex / insular cortex, left* | 99 | 4.5 | -34 | 22 | -8 |
| *superior frontal gyrus, right* | 88 | 4.01 | 4 | 28 | 58 |

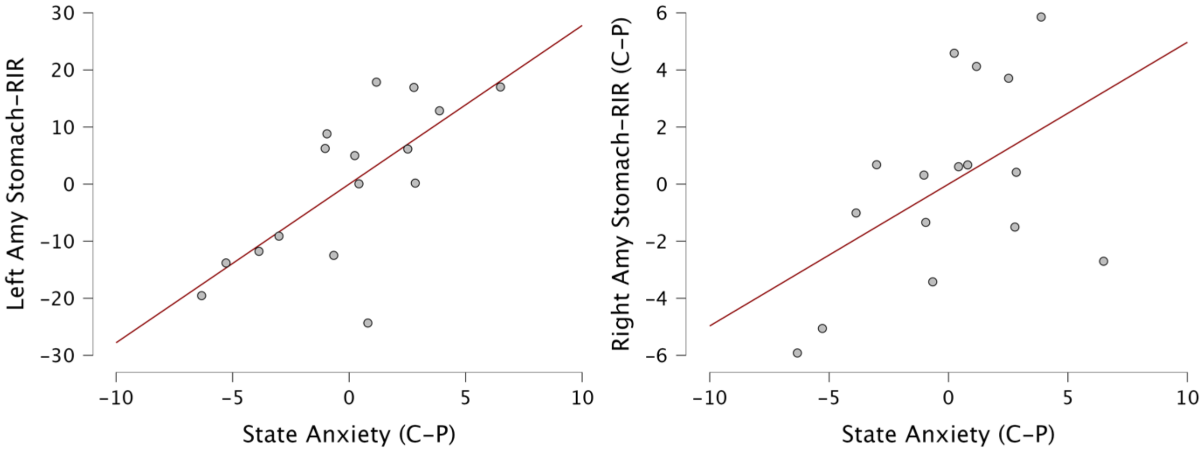

**Figure S7. Association Between CITALOPRAM Effect on State Anxiety and Neural Response to Stomach Focus (stomach-RIR) in a Robust Dataset.** Partial residual regression plots, after controlling for treatment order, change of nausea, change of headache and change of dizziness. C-P is CITALOPRAM – PLACEBO. Right Amy = right amygdalar cluster, Left Amy = left amygdalar cluster. Left amygdala cluster (b=2.7, t(10)=3.09, $p$ = .011), right amygdala cluster (b=.50, t(10)=1.8, $p$ = .096).

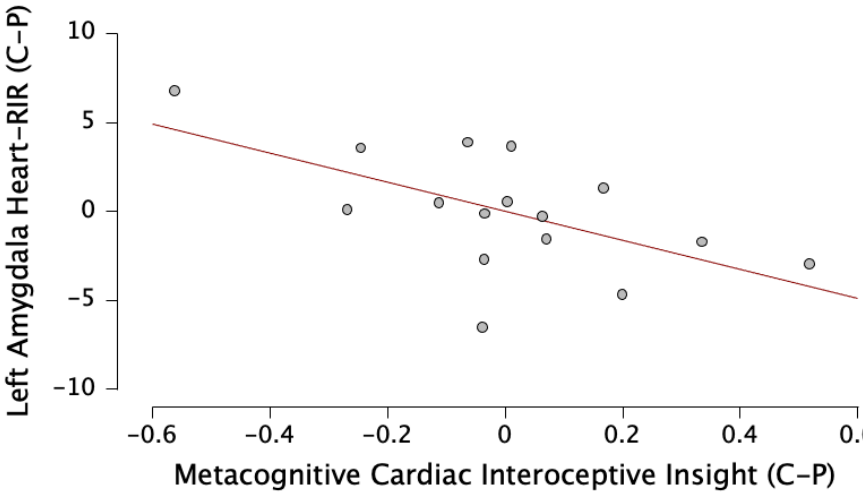

**Figure S8. Association Between CITALOPRAM Effect on Metacognitive Cardiac Interoceptive Insight and Neural Response to Heart Focus (heart-RIR) in a Robust Dataset.** Partial residual regression plots, after controlling for treatment order and change of heart rate. C-P is CITALOPRAM – PLACEBO. (b=-8.15, t(12)=-2.5, r = -.59, $p$ = .025)

**Table S5: fMRI Effect Clusters of STAI Association with Heart-RIR.** Contrast, region (Oxford Harvard Cortical and Subcortical Atlas), number of voxels in cluster, peak voxel z score and peak coordinates (x,y,z) in MNI space. Cluster-corrected analysis ($Z>3.1$, $p<0.05$).

| ***Anatomy*** | ***Voxels*** | ***Z MAX*** | ***X*** | ***Y*** | ***Z*** |
|---|---|---|---|---|---|
| *Superior Frontal Gyrus* | 721 | 5.97 | 10 | 36 | 54 |
| *Frontal Pole* | 635 | 5.38 | 42 | 44 | -4 |
| *Cerebellum* | 338 | 5.03 | 0 | -76 | -24 |
| *Frontal Orbital Cortex / Anterior Insular Cortex* | 316 | 5.13 | 44 | 30 | -12 |
| *Supramarginal Gyrus* | 306 | 5.61 | 50 | -42 | 56 |
| *Middle Temporal Gyrus* | 270 | 4.74 | 66 | -30 | -4 |
| *Cerebellum* | 266 | 5 | -44 | -64 | -48 |
| *Cerebellum* | 114 | 4.48 | -4 | -42 | -18 |
| *Cerebellum* | 101 | 4.58 | 24 | -34 | -26 |
